# Psychosocial Risk Factors for Injury in Performing Artists: A Scoping Review of Screening and Predictive Tools

**DOI:** 10.1101/2025.01.19.25320784

**Authors:** Róisín Cahalan, Caoimhe Barry Walsh, Orfhlaith Ni Bhriain, Breandán de Gallaí, Hannah Fahey de Brún, Michele Pye, Rose Schmieg

**Affiliations:** School of Allied Health, Physical Activity for Health Research Centre, University of Limerick, Ireland; School of Allied Health, University of Limerick, Ireland; Irish World Academy of Music and Dance, University of Limerick, Ireland; Shenandoah University, Division of Athletic Training, Shenandoah University, Virginia, United States; Howard University, Washington DC, United States

**Keywords:** Injury, Performing artists, Psychosocial risk, Screening/prediction

## Abstract

The performing arts are a broad church and include artists from various fields including dance, music and circus. Despite obvious differences in disciplines, much is shared in terms of an elevated risk of injury, and a common identity as a performer and artist. Injury screening protocols are imperative to mitigate injury risk. However, current programmes focus overwhelmingly on physical risks for injury and fail to appropriately consider psychosocial drivers injury affecting this cohort. This scoping review will synthesize the information on tools used to evaluate psychosocial factors related to injury in non-recreational performing artists, to identify robust instruments for use in future screening protocols. The scoping review will conform to Joanna Briggs Institute (JBI) Evidence Synthesis guidelines. Multiple databases relating to health, medicine, kinesiology, sport and dance will be searched for relevant articles. Two independent reviewers will conduct title and abstract screening followed by full-text screening. Data charting will be completing using a modified standardised form. Descriptive results will be reported using tabular and graphical media. The published scoping review will be disseminated to relevant stakeholders in the performing arts as well as to clinicians working with these artists. The resulting outputs will be in the form of both peer-reviewed and non-peer reviewed publications (e.g. blog posts, academic publications and conference presentations to reach key stakeholders such as performing artists and their support teams). An infographic of key findings will be developed and shared on social media platforms as appropriate. Ethical approval was not required for this study.

**What is already known on this topic:** – Although injury in the performing arts is extremely prevalent with implications for the biopsychological and financial wellbeing of the artists, there are few screening instruments which consider drivers of injury apart from physical factors.

**What this study adds:** – this study will identify psychosocial screening and predictive tools used to identify injury in performing artists, and the psychometric properties and utility of each.

**How this study might affect research, practice or policy:** – the outputs of this scoping review will raise stakeholder awareness of the psychosocial drivers of injury in performing artists. It will also identify a range of robust instruments that will aid in the holistic identification of those artists at risk of injury.

## INTRODUCTION/BACKGROUND

The performing arts is a broad church that may be defined as “art forms that are expressed by individuals or groups that involve performance through multi-sensory experiences, which may include, but need not be limited to, dance, song, music, theatre, and digital or electronic productions” (https://www.lawinsider.com/dictionary/performing-arts). Although predominantly aesthetic activities, many performing artists exist at the nexus between the artist and athlete. Significant physical and psychological ability and reserves are required to withstand the rigours of the highly repetitive nature of performance training and the threat of pain and injury.^1^

The incidence of injury across various fields in the performing arts is well documented. A recent review of dancers from various genres and at different levels (recreational, student elite and professional dancers) reported that injury in dance remains a concerning issue with a paucity of high-level evidence and inconsistency in research methods.^2^ Similarly in musicians, high levels of pain, weakness and other musculoskeletal disorders have been recorded.^3,4^ Phonotrauma in singers is largely understood to stem from cumulative vocal fold tissue damage and/or response to persistent tissue inflammation.^5^ It is estimated that up to 29% of attendees at voice clinics in the United States are professional singers, even though they represent less than 1% of the national work force.^6^

Interestingly, there are considerable overlaps in many of the identified causes of pain and injury across the performing arts. Much of the research in this area focuses on physical causes of injury, with factors including overtraining, excessive load and under-recovery identified in studies across music, song and dance respectively.^7-9^ A study exploring injury in dancers and musicians identified a host of biomechanical issues, poor ergonomics and suboptimal technique as key factors driving injury in both groups.^10^ Prior injury is also noted as an important risk factor for future injury across many areas of the performing arts.^11-13^

There is also a growing appreciation of the association between psychosocial issues and injury across various fields in the performing arts. Music performance anxiety and high levels of stress were found to be associated with an increased level of musculoskeletal disorders in a systematic review of professional and pre-professional instrumentalists.^12^ Emotional exhaustion, poor self-efficacy and fatigue were related to an increased injury risk in a cohort of circus artists^14^ while in singers an association between personality traits/facets related to happiness, dominance and caution, and phonotrauma were reportet.^8^ A multitude of factors including stress, psychological distress, disordered eating, poor coping, suboptimal sleep, personality, and social support have also been found to be associated with increased injury risk or adverse injury outcomes in dancers.^15^

In addition to shared injury experiences, performing artists have much in common regarding the identity of the artist, and attitudes to performance when ill or injured. Continuing to perform when in pain or injured is a common finding in dancers^16^, musicians^17^ and singers.^18^ These authors cite reasons that range from the more benign, such as a mild injury that it likely to resolve itself, to more sinister motivations including stigma associated with being injured, a culture of concealment, and fear of losing roles, work and status. Additionally, the experience of being seriously injured can be devastating for the performing artist, whose identity is intimately entwined with their art.^19,20^ The extent to which the performing artist experiences these challenges to their identity, and repercussions therein are known to be impacted by many factors including personality traits, coping strategies, and quality of social support.^21^

While the increasing focus on psychosocial drivers of pain and injury is encouraging, the overwhelming amount of research in this area is still devoted to exploring the pantheon of physical risk factors for injury, as well as offering guidance in addressing these issues.^1^ Recent efforts to develop a holistic screening tool for performing artists including physical and psychological evaluations specific to individual cohorts are encouraging.^22^ However, the emphasis remains on the physical experience of the artist with non-physical factors often siloed into individual fields. It is our contention that, despite the heterogeneous physical demands across the spectrum of performing artists, the research would indicate much that is shared in terms of identity, psychosocial risk for, and consequence of, injury. Failing to consider the shared psychosocial drivers of pain and injury therefore undermines the optimisation of holistic health and wellness in these performers.

This scoping review aims to systematically map and evaluate existing questionnaires, surveys and other relevant tools that evaluate, screen for or otherwise assess psychosocial risks for injury in non-recreational performing artists. Psychometric variables (specificity, sensitivity, validity, reliability as available) of each tool will be recorded. Ultimately, this protocol will inform future research and contribute to the development of standardized, evidence-based screening protocols to enhance holistic injury prevention in performing artists.

## METHODS AND ANALYSIS

This scoping review will be conducted in accordance with the Joanna Briggs Institute (JBI) Evidence Synthesis guidelines^23^ and Preferred Reporting Items for Systematic Reviews and Meta-Analyses extension for Scoping Reviews (PRISMA-SCr).^24^ Building on a prior framework and methodological guidance, this approach facilitates enhanced development and reporting of appropriate objectives and comprehensive search strategies. Precision of the topic was further determined through the use of concept framework focus the title, aims and objectives of the review. This facilitated a comprehensive search strategy, enhanced transparency and rigour of reporting and synthesis and presentation of findings (Table 1).

**Table 1:**
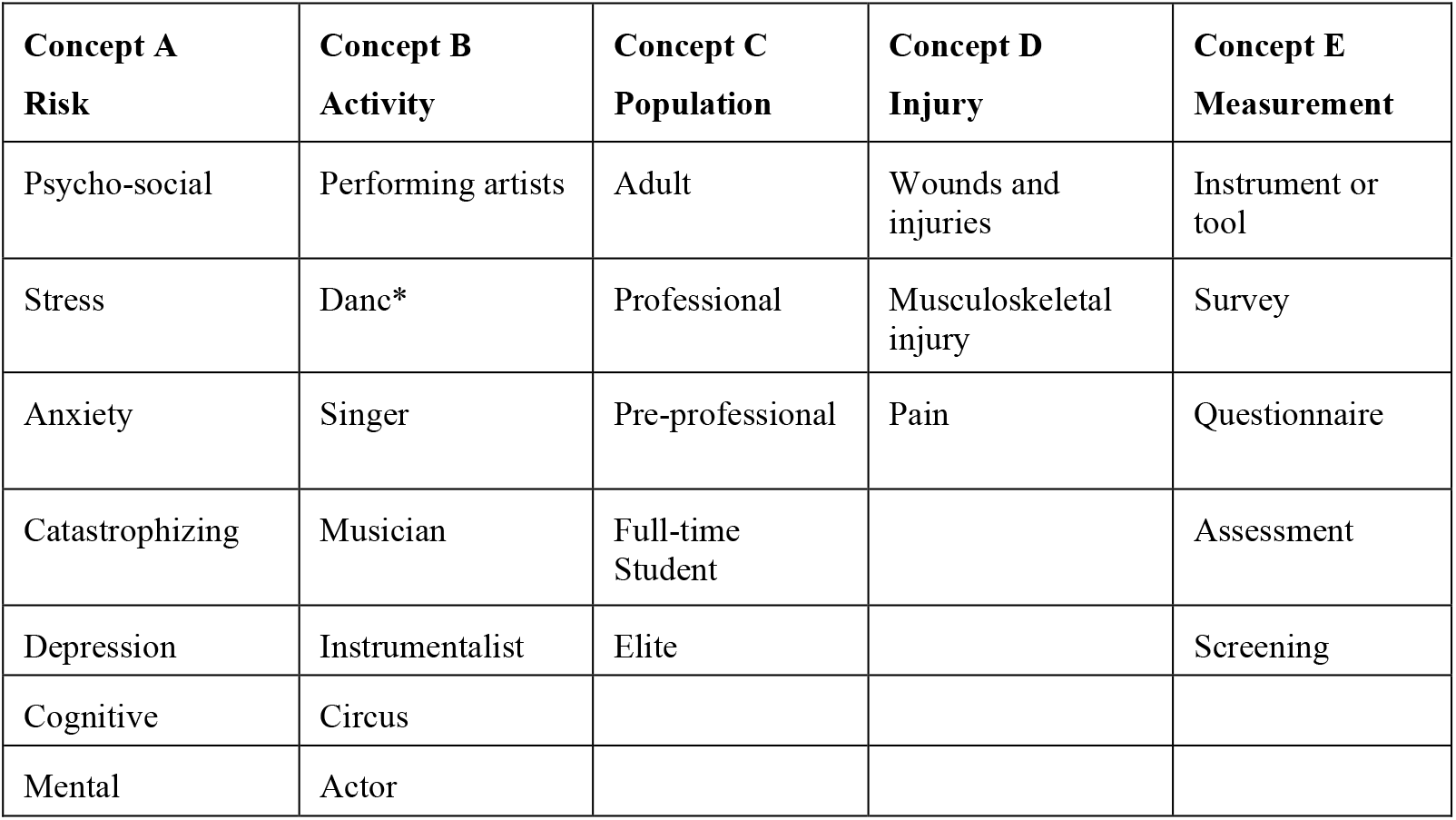
Key concepts informing search strategy

### Inclusion and exclusion criteria

The following criteria will be applied to the search in this scoping review:

### Inclusion criteria

- Original peer-reviewed research, clinical practice guidelines reporting assessment or evaluation tools (including surveys, questionnaires) of psychosocial factors associated with injury, pain or other physical ailment in performing artists. Pertinent systematic reviews in the area will be screened for studies meeting the inclusion criteria.
- Studies involving adult (18 years and over) dancers, musicians, singers, circus artists, or other physical performance (e.g. theatre actors).
- English language, human research studies published from the date of inception of the database.
- Non-recreational performing artist, including professional, pre-professional, full-time University (or equivalent) performing arts students, or otherwise described elite performers will be included. Studies of mixed (recreational and non-recreational) artists will be included only data pertaining to non-recreational performing artists may be extracted.

### Exclusion criteria

- Studies including tools which assess/measure solely physical factors including load, unless psychosocial factors can be individually extracted.
- Tool is not used to assess a relationship with pain/injury/other physical ailment.
- Qualitative (non-tool based) reports of psychosocial issues/risks.
- Studies of recreational performing artists.
- Studies of performing artists under the age of 18 years.

### Information Sources and Search Strategy

The search strategy has been developed by the research team, who share a wealth of clinical and research experience in various fields in the performing arts. The search strategy aimed to locate published primary studies. An initial limited search of Web of Science and CINAHL Ultimate (EBSCO) was undertaken to identify articles on the topic. The text words contained in the titles and abstracts of relevant articles, and the index terms used to describe the articles were used to develop a full search strategy for MEDLINE (EBSCO) (Appendix I, supplementary material).

The search strategy, including all identified keywords and index terms, was adapted for each database from inception; Databases are selected based on their relevance to the research topics of health, medicine, kinesiology, sport and dance. These databases include: Web of Science; EMBASE; Cochrane Database of Systematic Reviews; EBSCO (CINAHL Ultimate, MEDLINE, SPORTDiscus, PsycINFO); PubMed; Elsevier (ScienceDirect; Scopus); ProQuest Performing Arts Periodical Database, Dissertations); SAGE; JSTOR; and PEDro: the Physiotherapy Evidence Database. Grey literature will also be searched for pertinent articles (OpenGrey and GreyLit.org).

### Study Selection and Screening

The reference lists of articles included in the review will be screened for additional papers. Studies will be limited to those published in English and on human subjects. Following the search, all identified records will be collated and uploaded into Endnote X9.3.3(Clarivate Analytics, PA, USA) and duplicates will be removed. Following a pilot test, titles and abstracts will then be screened by two independent reviewers (RC & CBW) for assessment against the inclusion and exclusion criteria for the review. Potentially relevant papers will be retrieved in full and their citation details imported into the Covidence reference management system. (Covidence; Covidence Melbourne, Australia). The full text of selected citations will be assessed in detail against the inclusion and exclusion criteria by two independent reviewers (RC & CBW). Reasons for exclusion of full-text papers will be recorded and reported in the scoping review. Any disagreements that arise between the reviewers at each stage of the selection process will be resolved through discussion or with a third reviewer (RS). Authors of papers will be contacted to request missing or additional data, where required. Full texts of selected papers will be hand searched for additional relevant papers referred to in the appendices. If access to missing data is not possible these papers will be excluded. The results of the search will be reported in full in the final scoping review and presented in a Preferred Reporting Items for Systematic Reviews and Meta-analyses for Scoping Reviews (PRISMA-ScR) flow diagram.^24^

### Data Charting/Collection/Extraction

Data charting, collection, and extraction for this scoping review will follow a systematic and transparent process to ensure comprehensive capture of relevant information. A standardized data extraction form will be developed, detailing key information from eligible studies. This will include study characteristics (author, year, country of origin), characteristics of tools (name, version of tool, purpose, range of the scores/points, cut-off points where available, administration method, and the duration of administration), study design, study population (profession, sex, age), settings, the psychometric performance of tools where available (including sensitivity, specificity, reliability and validity). Data will be charted in a systematic matrix or table for easy comparison, summarizing key domains to inform the development of a future tool for return to dance. The details of “friend studies” will be included only if they add additional, relevant data to that contained in the related study. A risk of bias evaluation will be completed by author RC, using the Downs and Black Checklist.^25^ This tool assesses the methodological quality of both randomized and non-randomized studies. It focuses on bias across a broad range of domains such as reporting, external validity, and statistical analysis.

### Synthesis and Presentation of Results

A PRISMA flow diagram will document the selection process, from database searching and article screening to the final selection of studies for inclusion in the review. Selected data will be extracted and presented in a table with fields including study details (author/year), population, nature of the tool, nature of injury, domains included, purpose of the tool, administration details, and any associated psychometric properties where available (Appendix 2, Supplementary material). Data synthesis will involve categorizing and summarizing the key findings of the included studies, such as the types of instruments used, domain properties, populations studied, and any identified gaps or challenges in establishing readiness for return to practice. Descriptive analysis with basic coding will be used to present the type and frequency of common domains used across studies, which will be presented in table or graphical format.

## Discussion and Dissemination

The performing arts are a broad church and includes artists from a range of different fields. However, much is shared in terms of an elevated risk of injury, and a common identity as a performer and artist. Mitigating the risk of injury in this cohort is key to supporting holistic wellness, quality of life and financial security. Screening programmes for factors exposing performers to injury are therefore of great importance. However, current programmes focus overwhelmingly on physical risks for injury and fail to appropriately consider the many psychosocial drivers of pain and injury in this cohort. This scoping review will synthesize the information on tools used to evaluate psychosocial factors related to injury in non-recreational performing artists, in order to identify the most robust instruments for use in future screening protocols. The results of this scoping review will inform future work in developing high quality, generic tools to be integrated into screening tools for performing artists from a broad array of disciplines.

The results of this scoping review will be disseminated in both academic and performing arts fora. An academic paper will be submitted for peer reviewed publication, and to appropriate conference(s). A lay summary will also be made available to a non-academic audience and distributed to the professional and education performing arts communities. These include performing arts conservatoires, companies and higher education institutions. Once published the results of the study will be summarised and shared in plain language to dancers in digital format (X, Instagram) and through the Universities of the authors and affiliated performing arts organisations.

## Data Availability

All data produced in the present work are contained in the manuscript

