## Supplemental material for "Psychosocial Risk Factors for Injury in Performing Artists: A Scoping Review of Screening and Predictive Tools"

**Supplementary material**

### Appendix 1: Search strategy

Limited to adults and studies in the English language

("psychosocial factors"[Title/Abstract] OR "mental health"[Title/Abstract] OR "anxiety"[Title/Abstract] OR "depression"[Title/Abstract] OR "catastrophizing"[Title/Abstract] OR "catastrophising"[Title/Abstract] OR "psychological stress"[Title/Abstract] OR "burnout"[Title/Abstract] OR "cognitive factors"[Title/Abstract] OR "stress"[Title/Abstract])

AND

("performing artists"[Title/Abstract] OR "danc*"[Title/Abstract] OR "musician*"[Title/Abstract] OR "instrumentalist*"[Title/Abstract] OR singer*[Title/Abstract] OR vocalist*[Title/Abstract] OR actor*[Title/Abstract] OR "circus performer*"[Title/Abstract] OR "circus arts"[Title/Abstract] OR "performing arts"[Title/Abstract] OR "occupational performer*"[Title/Abstract]

AND

"elite*"[Title/Abstract] OR "professional*"[Title/Abstract] OR "pre-professional *"[Title/Abstract] OR "non-recreational"[Title/Abstract])

AND

("pain"[Title/Abstract] OR "injury"[Title/Abstract] OR "musculoskeletal pain"[Title/Abstract] OR "chronic pain"[Title/Abstract] OR "musculoskeletal disorders"[Title/Abstract] OR "injury prevention"[Title/Abstract] OR "injury risk"[Title/Abstract] OR "workplace injury"[Title/Abstract])

AND

("screening tools" OR "predictor tools" OR "predictive tools" OR "questionnaire*" OR "outcome measures" OR "risk assessment tools" OR "assessment tools" OR "evaluation tools" OR "diagnostic tools" OR "measurement tools")

### Supplement 2: Extracted data table

| Study Characteristics | Study design | Population | Characteristics of Tool | Setting | Outcomes | Psychometric Properties |
|---|---|---|---|---|---|---|
| Author<br>Year<br>Country of Origin | Cross-sectional<br>RCT<br>Prospective | Profession<br>Sex<br>Age | Name, Version<br>Purpose<br>Scoring and cut-offs<br>Administration method<br>Duration | Clinic<br>Studio | Association risk factor and injury | Reliability<br>Validity<br>Sensitivity<br>Specificity. |
